# Gender and Geographic Diversity of the Decentralised Leadership of Global Oral Health Organisations

**DOI:** 10.1101/2025.06.09.25329297

**Authors:** Ratilal Lalloo

## Abstract

**Purpose:** The purpose of the paper is to analyse the gender and geographic diversity of the decentralised leadership of global oral health organisations.

**Methodology:** Publicly available data were accessed on the gender and location of the decentralised leadership across the International Association for Dental, Oral, and Craniofacial Research (IADR), World Dental Federation (FDI) and International Federation of Dental Hygienists (IFDH). Gender was allocated from photographs if available on the organisation websites and other online profiles, and using Genderize, an online allocation platform.

**Findings:** This analysis shows positive findings on gender diversity of the decentralised leadership, across the organisations. The most senior leadership roles (Presidents) are more likely to be male except for the IADR Scientific Groups and Networks (SGN) where more than half are female. Geographic diversity where relevant is less diverse, with the majority of leaders from high-income countries in North America and Europe. This is illustrated by the senior leadership roles in SGNs being almost exclusively from these same two continents.

**Originality/value:** The decentralised leadership is gender diverse, but senior roles and some groups are male dominated. Decentralised leaders are from high-income countries in the Global North. There is an urgent need to implement strategies to address equity, diversity and inclusivity in the decentralised leadership. A lack of diversity here means that central leadership will continue to lack diversity.

## Introduction

Equity, diversity and inclusivity in oral health have focused on leadership of academic institutions and research productivity. Oral health leadership reflects a dominance by men (Tiwari et al., 2019, Li et al., 2019, Weinstein et al., 2022, Bompolaki et al., 2022). Academic leadership in particular is dominated by men, with women making up about a third of dental researchers (Tiwari et al., 2019, Gangwani and Kolokythas, 2019).

A recent analysis of the diversity of the central leadership of global oral health organisations showed positive gender diversity trends but leaders were essentially from high-income countries in the Global North (Lalloo, 2024). The analysis showed that a third of the International Association for Dental, Oral, and Craniofacial Research (IADR) Board Directors (n=15) were female and 40% were from the American division. Of the World Dental Federation (FDI) Council members (n=15) more than half (53%) were female, however men held all senior roles. Two-thirds of Council members were from Europe and the USA and 42% of Standing Committee members were from Europe. Of the 100 IADR past presidents (1921-2023) only 11 have been female. Of the 47 FDI past presidents (1901-2023) only six have been female. Almost 90% of FDI past presidents are from high-income Global North countries. Across all global health organisations analysed central leadership from low- and middle-income countries, and more generally the Global South, is lacking.

Representation of the global workforce and population within oral health organisations, at both central and grassroot levels, is critical to ensure a global agenda and priorities. This paper aims to complement the previous work on the diversity of the centralised leadership and shed light on the current state of the gender and geographic diversity of the decentralised (grassroots) leadership of global oral health organisations. Grassroots leaders are potentially the future leaders at the central level. Enhancing diversity here can potentially enhance diversity at the central level.

## Methods

The gender and geographic diversity of the decentralised leadership of the International Association for Dental, Oral, and Craniofacial Research (IADR), World Dental Federation (FDI) and International Federation of Dental Hygienists (IFDH) were analysed from publicly available data on the organisations’ respective websites (https://www.iadr.org/, https://www.fdiworlddental.org/ and https://ifdh.org/). The data were collected across September 2024.The IADR leadership was analysed for Regions, Divisions and Sections (RDS) and Scientific Groups and Networks (SGN). The FDI leadership was analysed for membership of Task Teams, Working Groups and Expert Groups. The IFDH leadership was analysed for Committees.

Male or female was allocated to leaders from photographs on the organisation websites, other online profiles and by using a gender allocation from Genderize, an online software (https://genderize.io/). First names of the leaders were uploaded to the platform. The gender diversity was compared by roles, RDS, SGN and continent. The locations of leaders included RDS and continent.

## Results

### IADR

There are five IADR regions: Africa/Middle East; Asia/Pacific; Latin American; North American and Pan European. Of the 59 leaders across these regions, 41% are female (Table I). Of the four Presidents, one is female. Of the 37 members, 40% are female. By region, and with those with more than 10 leaders, 42% of leaders in the Africa/Middle East, 31% in Asia/Pacific and 45% in Pan Europe are female.

**Table I.**
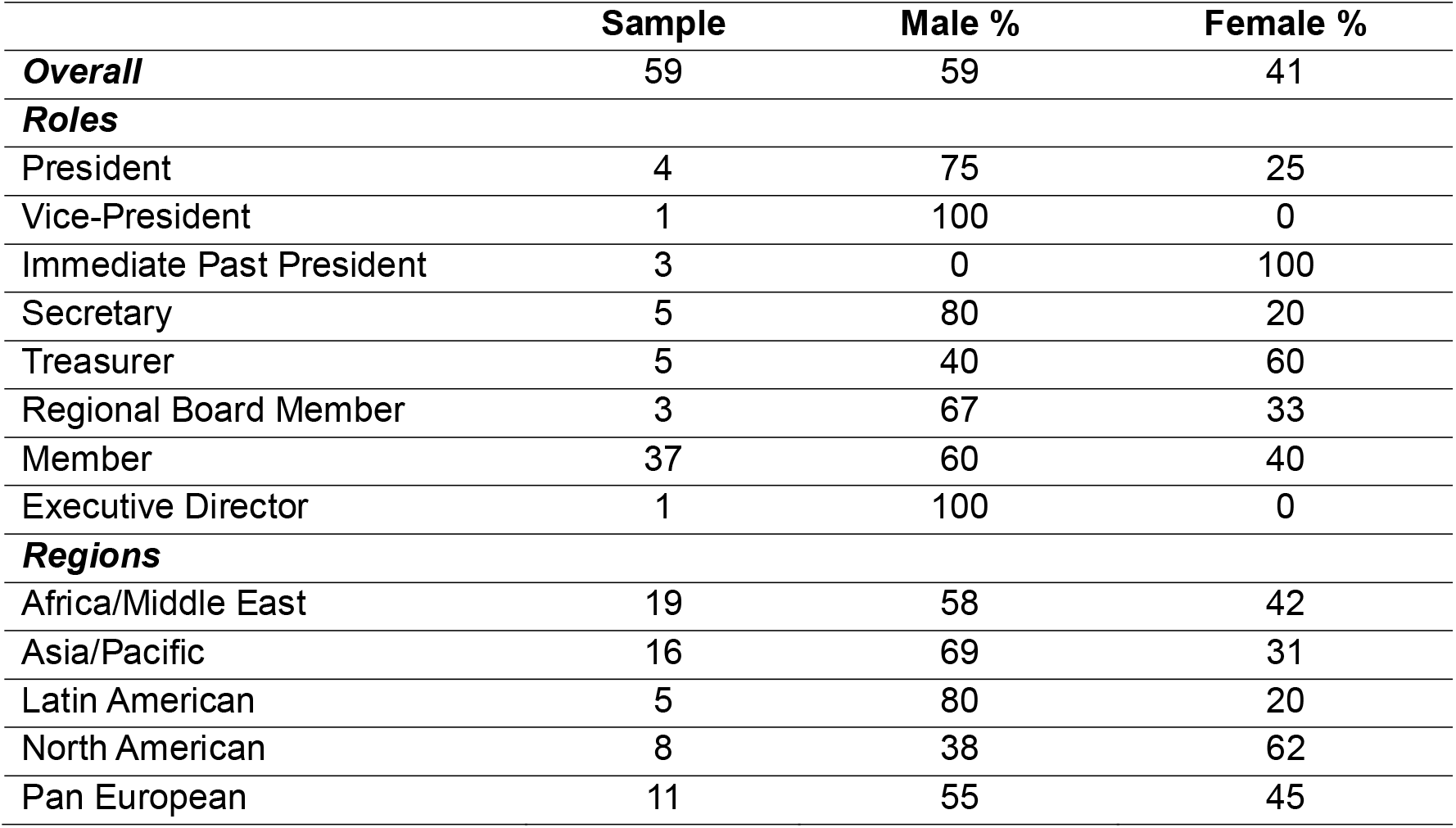
Gender Diversity of IADR Regional Leadership by Roles and Regions.

|  | **Sample** | **Male %** | **Female %** |
| --- | --- | --- | --- |
| ***Overall*** | 59 | 59 | 41 |
| ***Roles*** |  |  |  |
| President | 4 | 75 | 25 |
| Vice-President | 1 | 100 | 0 |
| Immediate Past President | 3 | 0 | 100 |
| Secretary | 5 | 80 | 20 |
| Treasurer | 5 | 40 | 60 |
| Regional Board Member | 3 | 67 | 33 |
| Member | 37 | 60 | 40 |
| Executive Director | 1 | 100 | 0 |
| ***Regions*** |  |  |  |
| Africa/Middle East | 19 | 58 | 42 |
| Asia/Pacific | 16 | 69 | 31 |
| Latin American | 5 | 80 | 20 |
| North American | 8 | 38 | 62 |
| Pan European | 11 | 55 | 45 |

There are 42 active (with members) IADR Divisions and Sections. Of the 262 leaders, 48% are female (Table II). Of the 42 Presidents 29% are female. Of the 26 Vice-Presidents 38% are female. More than half of Secretaries (53%) and Treasurers (61%) are female. The number of leaders by RDS are small but differences in gender diversity can be observed.

**Table II.** Gender Diversity of IADR Divisional Leadership by Roles and Divisions / Sections.

|  | **Sample** | **Male %** | **Female %** |
| --- | --- | --- | --- |
| ***Overall*** | 262 | 52 | 48 |
| ***Roles*** |  |  |  |
| President | 42 | 71 | 29 |
| President-Elect | 8 | 38 | 62 |
| Vice-President | 26 | 62 | 38 |
| Immediate Past President | 16 | 56 | 44 |
| Secretary | 34 | 47 | 53 |
| Treasurer | 31 | 39 | 61 |
| Councillor | 62 | 57 | 43 |
| Member | 19 | 21 | 79 |
| Other | 29 | 52 | 48 |
| ***Divisions / Sections*** |  |  |  |
| American | 15 | 33 | 67 |
| Australia/New Zealand | 17 | 53 | 47 |
| Argentine | 12 | 25 | 75 |
| Bolivian | 4 | 25 | 75 |
| Brazilian | 14 | 71 | 29 |
| British | 5 | 40 | 60 |
| Canadian | 7 | 43 | 57 |
| Caribbean | 4 | 50 | 50 |
| Chilean | 4 | 75 | 25 |
| Chinese | 13 | 77 | 23 |
| Colombian | 6 | 33 | 67 |
| Continental Europe | 12 | 42 | 58 |
| Costa Rican | 5 | 60 | 40 |
| East & Southern Africa | 6 | 33 | 67 |
| Ecuadorian | 1 | 100 | 0 |
| Egyptian | 3 | 100 | 0 |
| Guatemalan | 1 | 100 | 0 |
| Indian | 2 | 100 | 0 |
| Iranian | 4 | 100 | 0 |
| Iraqi | 6 | 50 | 50 |
| Irish | 3 | 33 | 67 |
| Israeli | 2 | 50 | 50 |
| Japanese | 4 | 75 | 25 |
| Jordanian | 6 | 67 | 33 |
| Korean | 9 | 78 | 22 |
| Kuwaiti | 9 | 56 | 44 |
| Lebanese | 4 | 100 | 0 |
| Libyan | 2 | 100 | 0 |
| Mexican | 2 | 0 | 100 |
| Nigerian | 3 | 67 | 33 |
| Pakistani | 9 | 78 | 22 |
| Palestinian | 7 | 57 | 53 |
| Peruvian | 2 | 0 | 100 |
| Qatar | 3 | 100 | 0 |
| Saudi Arabian | 4 | 25 | 75 |
| Scandinavian | 11 | 27 | 73 |
| South African | 10 | 10 | 90 |
| Southeast Asian | 11 | 55 | 45 |
| Tunisian | 3 | 0 | 100 |
| United Arab Emirates | 11 | 73 | 27 |
| Uruguayan | 4 | 75 | 25 |
| Venezuelan | 7 | 14 | 86 |

There are 36 SGNs, with 369 leaders and equal representation by gender (Table III). More than half (53%) of the 34 Presidents, less than half (45%) of the 33 President- Elects and 40% of the 35 Vice-Presidents are female. Of the 33 Secretaries / Treasurers 60% are female. Of the 37 Councillors 59% are female. Both by SGN and RDS differences in gender diversity can be observed. Of the 369 SGN leaders 71% are from four divisions; 159 (43%) are from the American, 43 (12%) from the Continental Europe, 39 (11%) from the British and 20 (5%) from the Brazilian divisions. Of the 369 three-quarters are from two continents, North America (n=176; 48%) and from Europe (n=100; 27%). In these respective divisions and continents the majority of leaders are female. By role, almost all of the 34 Presidents are from North America (n=20; 59%) and Europe (n=12; 35%). Of the 33 President-Elects, 13 (39%) are from North America, 10 (30%) from Europe and 8 (24%) from Asia. Supplementary Table I presents the percentage distribution of SGN roles by continent.

**Table III.** Gender Diversity of IADR Scientific Groups and Networks Leadership by Roles, Divisions / Sections and Continent.

|  | Sample | Male % | Female % |
| --- | --- | --- | --- |
| **Overall** | 369 | 50 | 50 |
| **Roles** |  |  |  |
| President | 34 | 47 | 53 |
| President-Elect | 33 | 55 | 45 |
| Vice-President | 35 | 60 | 40 |
| Immediate Past President | 32 | 69 | 31 |
| Secretary/Treasurer | 33 | 40 | 60 |
| Councillor | 37 | 41 | 59 |
| Program Chair | 52 | 60 | 40 |
| Proposal Coordinator | 35 | 43 | 57 |
| Focused Learning Session Organizer | 12 | 50 | 50 |
| Hands-on Workshop Organizer | 16 | 31 | 69 |
| Symposium Organizer | 18 | 44 | 56 |
| Other | 32 | 44 | 56 |
| ***Specialist Groups / Networks*** |  |  |  |
| Behavioral, Epidemiologic, and Health Services | 13 | 62 | 38 |
| Cariology | 16 | 25 | 75 |
| Clinical and Translational Science | 8 | 63 | 37 |
| Craniofacial Biology | 14 | 36 | 64 |
| Dental Anesthesiology and Special Care | 15 | 53 | 47 |
| Dental Materials | 18 | 61 | 39 |
| Diagnostic Sciences | 9 | 56 | 44 |
| Digital Dentistry | 6 | 83 | 17 |
| e-Oral Health | 11 | 36 | 64 |
| Education | 22 | 32 | 68 |
| Evidence-based Dentistry | 8 | 75 | 25 |
| Geriatric Dentistry | 7 | 71 | 29 |
| Global Oral Health Inequalities | 8 | 50 | 50 |
| Implantology | 8 | 100 | 0 |
| International Network of Orofacial Pain and Related Disorders Methodology | 7 | 29 | 71 |
| Lasers & Bio-photonics | 13 | 84 | 16 |
| Microbiology & Immunology | 10 | 20 | 80 |
| Mineralized Tissues | 7 | 71 | 29 |
| Minimally Invasive Dentistry | 12 | 42 | 58 |
| Network of Practice-based | 1 | 100 | 0 |
| Neuroscience | 7 | 100 | 0 |
| Nutrition | 8 | 38 | 62 |
| Oral & Maxillofacial Surgery | 9 | 56 | 44 |
| Oral Health | 8 | 13 | 87 |
| Oral Malodor | 7 | 14 | 86 |
| Oral Medicine & Pathology | 7 | 71 | 29 |
| Orthodontics | 15 | 20 | 80 |
| Pediatric Oral Health | 9 | 44 | 56 |
| Periodontal | 15 | 60 | 40 |
| Pharmacology, Therapeutics, Toxicology | 3 | 33 | 67 |
| Prosthodontics | 10 | 70 | 30 |
| Pulp Biology and Regeneration | 13 | 62 | 38 |
| Salivary | 11 | 55 | 45 |
| Stem Cell Biology | 7 | 57 | 43 |
| Student Training & Research | 15 | 60 | 40 |
| Women in Science | 12 | - | 100 |
| ***Divisions / Sections*** |  |  |  |
| American | 159 | 46 | 54 |
| Australia/New Zealand | 14 | 29 | 71 |
| Argentine | 2 | 100 | 0 |
| Bolivian | - | - | - |
| Brazilian | 20 | 50 | 50 |
| British | 39 | 39 | 61 |
| Canadian | 17 | 59 | 41 |
| Caribbean | - | - | - |
| Chilean | 1 | 0 | 100 |
| Chinese | 4 | 100 | 0 |
| Colombian | - | - | - |
| Continental Europe | 43 | 42 | 58 |
| Costa Rican | - | - | - |
| East & Southern Africa | 1 | 0 | 100 |
| Ecuadorian | - | - | - |
| Egyptian | 3 | 0 | 100 |
| Guatemalan | - | - | - |
| Indian | 9 | 89 | 11 |
| Iranian | - | - | - |
| Iraqi | - | - | - |
| Irish | 5 | 60 | 40 |
| Israeli | 3 | 100 | 0 |
| Japanese | 6 | 100 | 0 |
| Jordanian | - | - | - |
| Korean | 1 | 100 | 0 |
| Kuwaiti | - | - | - |
| Lebanese | - | - | - |
| Libyan | - | - | - |
| Mexican | 2 | 100 | 0 |
| Nigerian | 1 | 0 | 100 |
| Pakistani | 2 | 100 | 0 |
| Palestinian | - | - | - |
| Peruvian | - | - | - |
| Qatar | 1 | 100 | 0 |
| Saudi Arabian | 1 | 100 | 0 |
| Scandinavian | 13 | 46 | 54 |
| South African | 1 | 0 | 100 |
| Southeast Asian | 19 | 68 | 32 |
| Tunisian | - | - | - |
| United Arab Emirates | 2 | 100 | 0 |
| Uruguayan | - | - | - |
| Venezuelan | - | - | - |
| **Continents** |  |  |  |
| Africa | 6 | 0 | 100 |
| Asia | 41 | 83 | 17 |
| Central America | 2 | 100 | 0 |
| Europe | 100 | 42 | 58 |
| Middle East | 7 | 100 | 0 |
| North America | 176 | 47 | 53 |
| Oceania | 14 | 29 | 71 |
| South America | 23 | 52 | 48 |

### FDI

There are 16 Task Teams, four Working Groups and one Expert Groups, with a total of 97 leaders, of whom 46% are female (Table IV). By role, 23% of the 22 Presidents and 53% of the 75 members are female. Of the 97 leaders, almost three-quarters are from Europe (n=43; 44%), Asia (n=17; 18%) and North America (n=12; 12%).

**Table IV.**
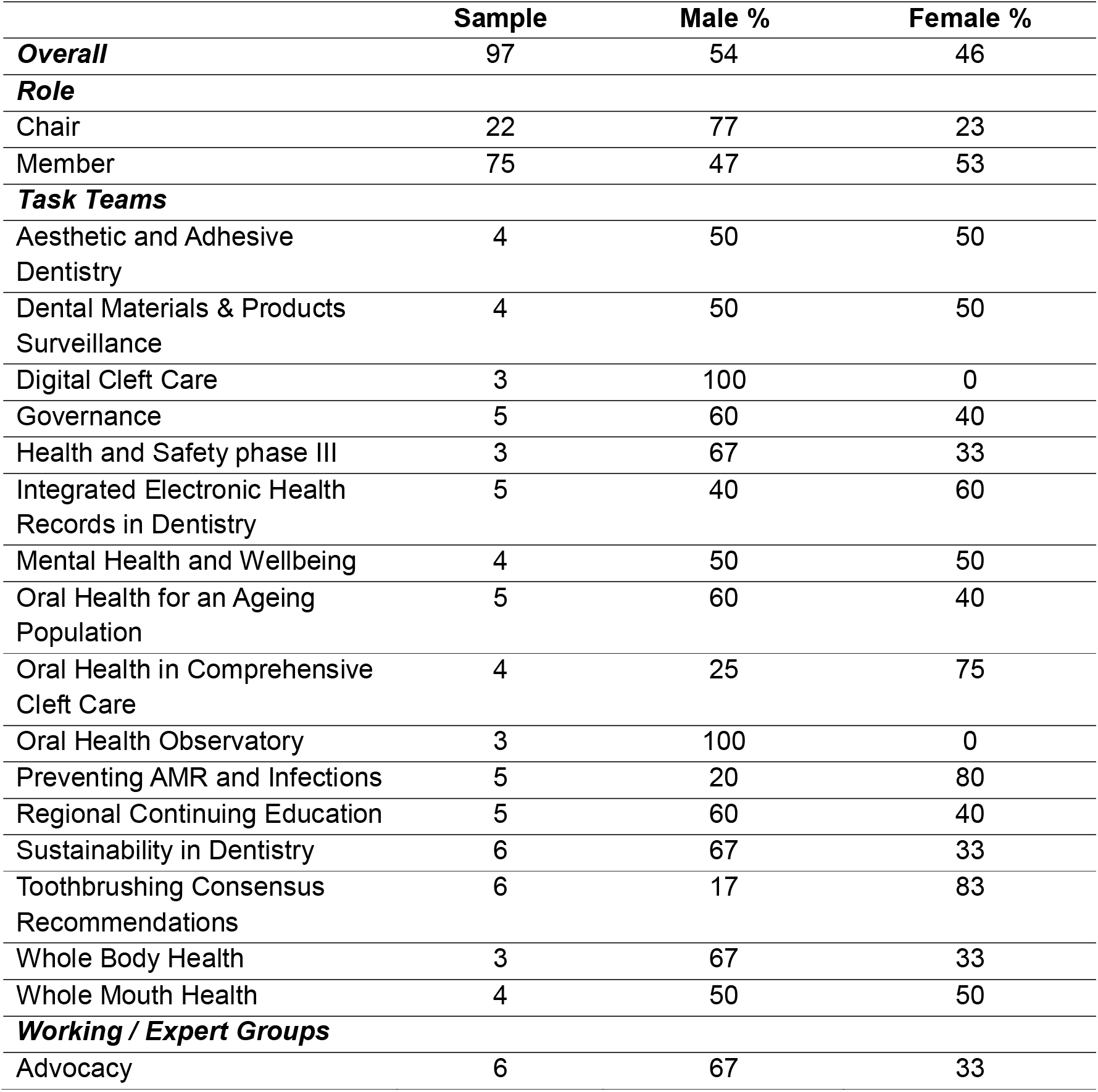
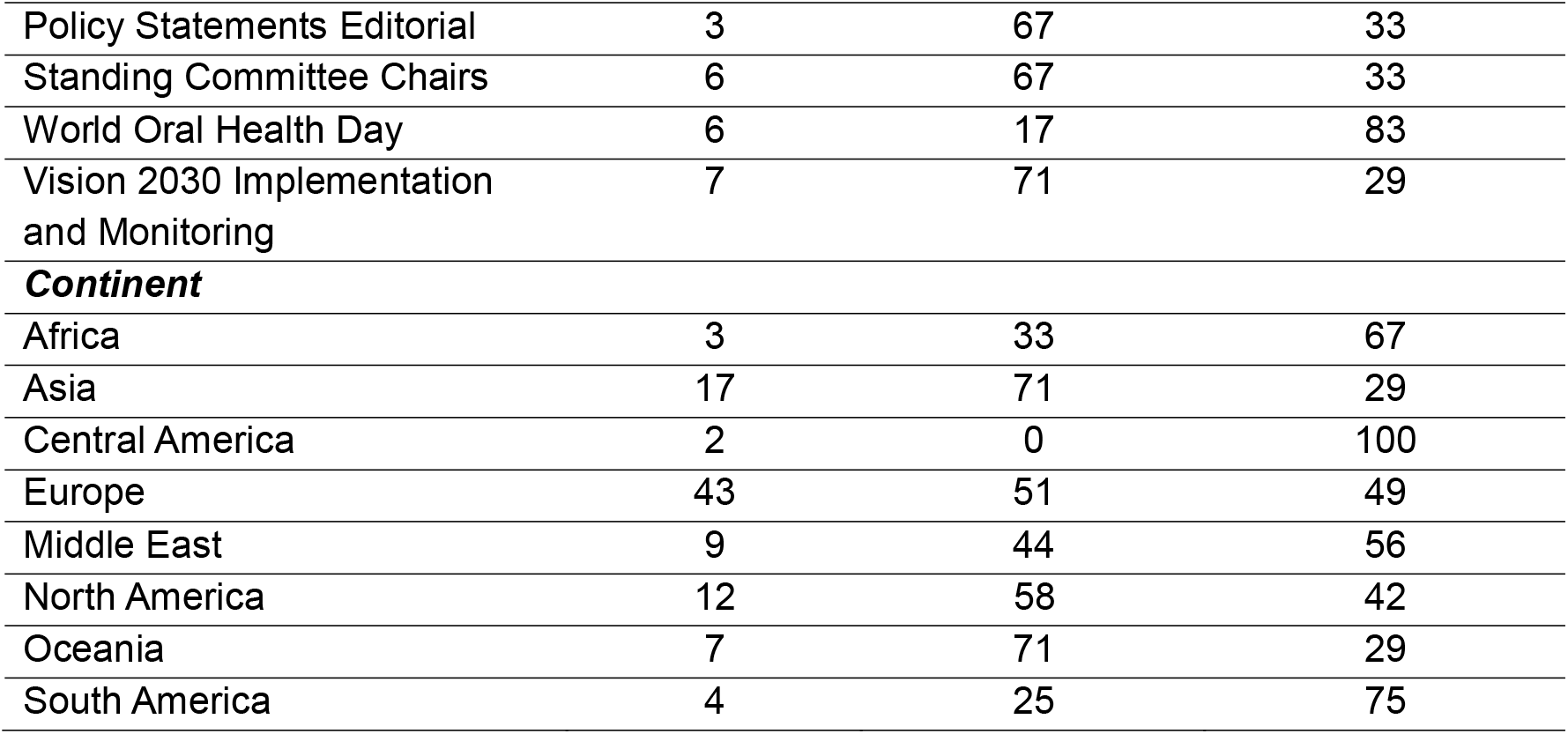
FDI Leadership by Roles, Task Teams, Working / Expert Groups and Continent.

### IFDH

There are seven IFDH Committees, and the website only lists the chairs or co-chairs. One committee chair position is not listed. All the 11 chairs are female. The chairs are from Canada (2); Netherlands; Lithuania; United Kingdom; United States (2); Sweden; Ireland; South Africa and Israel.

## Discussion

Addressing gender and geographic diversity in global oral health organisations is important to ensure the leadership reflects the changing oral health workforce and global society. The diversity needs to be reflected across the entire organisation, not only the central leadership. This analysis shows positive findings on gender diversity of the decentralised leadership, across IADR RDS and SGNs, as well FDI task teams, working and expert groups. However the most senior leadership roles (for example Presidents) are more likely to be male except for SGNs where more than half are female. Geographic diversity where relevant (at the Regions and Divisions this is not relevant as leaders come from these geographic regions) in the IADR SGNs and FDI is less diverse, with the majority of leaders from high-income countries in North America and Europe. This is also further illustrated by the senior leadership roles in SGNs being almost exclusively from these same two continents. Of the 34 Presidents, 94% are from North America and Europe. The President-Elect diversity is better in that a quarter are from Asia. This significant skew of the decentralised leadership to high-income countries needs to be urgently addressed, to reflect the global workforce and population. Not having a global decentralised leadership means that the central leadership will also lack geographic diversity. The decentralised leadership, especially in the senior leadership roles, will be a pathway to the central leadership. If the decentralised leadership is chosen by popular vote, then large divisions such as the America division will continue to dominate these positions. Membership distribution from the 2023 annual report shows about a third of the IADR members are from the American, 9% from the Continental Europe, 8% from the Japanese and 7% from the Chinese divisions.

Strategies to enhance diversity are widely available, including in publications specific to oral health (Smith and Sinkford, 2022, Garcia et al., 2020, D’Silva et al., 2019, Lalloo, 2022, Cain et al., 2022, Tiwari et al., 2019, Li et al., 2019). The IADR in March 2024 adopted a Diversity, Equity, Inclusion, Accessibility, and Belonging Statement. This statement includes the following: “*IADR expects all of its Divisions and Sections as well as Scientific Groups and Networks to strive towards these IADR DEIAB principles in everyday practice in furtherance of the IADR Mission*.*”* The American Association for Dental, Oral, and Craniofacial Research (AADOCR) recently published details and outcomes of a mentoring program (MIND the Future) to support *“the development of a diverse cadre of dental, oral, and craniofacial researchers”* (Herren et al., 2024). A core value of the FDI is *“Culture of inclusiveness: We deliberately and meaningfully engage and seek representation from the diverse range of oral health professionals and the communities and individuals they serve. This is paramount to achieving our mission*.*”* Strategies to address equity, diversity and inclusivity of the leadership need to be implemented across all levels of global oral health organisations.

Gender was only categorised as male and female, it was not possible to identify persons as non-binary and gender nonconforming. Other equity indicators such as ethnicity, race, age, socio-economic status and language and their interactions were not analysed (Muirhead et al., 2020). A diverse leadership at all levels has the enhanced potential to effectively address the complex global challenges facing the oral health profession and community. While this analysis is focussed on gender and geographic location, it is critical that organisations adopt an intersectionality lens when reflecting on their global membership and leadership at all levels of the organisation. Succession planning for decentralised leadership roles must take into account equity, diversity and inclusivity, and ensure a truly global representation.

## Data Availability

All data analysed are publicly available, on the websites of the global oral health organisations and on individual online profiles.

## Ethics statement

Data reported in this study are publicly available at the respective organisations’ websites or personal profiles of the leadership team members.

---

**Supplementary Table 1.** Percentage Distribution of Scientific Groups and Networks Leadership Roles by Continent.

| **Role** | **Sample** | **Africa** | **Asia** | **Central**<br>**America** | **Europe** | **Middle**<br>**East** | **North**<br>**America** | **Oceania** | **South**<br>**America** |
| --- | --- | --- | --- | --- | --- | --- | --- | --- | --- |
| President | 34 | 0 | 0 | 0 | 35 | 0 | 59 | 0 | 6 |
| President-Elect | 33 | 0 | 24 | 0 | 30 | 0 | 39 | 3 | 3 |
| Vice-President | 35 | 0 | 11 | 0 | 17 | 6 | 49 | 11 | 6 |
| Secretary / Treasurer | 33 | 6 | 21 | 0 | 24 | 0 | 33 | 9 | 6 |
| Program Chair | 52 | 0 | 10 | 2 | 27 | 0 | 48 | 2 | 12 |
| Councillor | 37 | 3 | 3 | 3 | 19 | 5 | 60 | 3 | 5 |
| Proposal Coordinator | 35 | 0 | 11 | 0 | 29 | 3 | 51 | 0 | 6 |
| Symposium Organizer | 18 | 0 | 0 | 0 | 50 | 0 | 50 | 0 | 0 |
| Focused Learning Session Organizer | 12 | 0 | 8 | 0 | 42 | 8 | 33 | 0 | 8 |
| Hands-on Workshop Organizer | 16 | 0 | 6 | 0 | 38 | 0 | 44 | 6 | 6 |
| Immediate Past President | 32 | 3 | 22 | 0 | 13 | 0 | 53 | 6 | 3 |
| Other | 32 | 6 | 9 | 0 | 28 | 3 | 41 | 3 | 9 |

